# Demonstrating the Consequences of Learning Missingness Patterns in Early Warning Systems for Preventative Health Care: A Novel Simulation and Solution

**DOI:** 10.1101/2020.06.05.20123323

**Authors:** Christopher E. Gillies, Daniel F. Taylor, Brandon C. Cummings, Sardar Ansari, Fadi Islim, Steve L. Kronick, Richard P. Medlin, Kevin R. Ward

## Abstract

When using tree-based methods to develop predictive analytics and early warning systems for preventive healthcare, it is important to use an appropriate imputation method to prevent learning the missingness pattern. To demonstrate this, we developed a novel simulation that generated synthetic electronic health record data using a variational autoencoder with a custom loss function, which took into account the high missing rate of electronic health data. We showed that when tree-based methods learn missingness patterns (correlated with adverse events) in electronic health record data, this leads to decreased performance if the system is used in a new setting that has different missingness patterns. Performance is worst in this scenario when the missing rate between those with and without an adverse event is the greatest. We found that randomized and Bayesian regression imputation methods mitigate the issue of learning the missingness pattern for tree-based methods. We used this information to build a novel early warning system for predicting patient deterioration in general wards and telemetry units: PICTURE (**P**redicting **I**ntensive **C**are **T**ransfers and other **U**nfo**R**eseen **E**vents). To develop, tune, and test PICTURE, we used labs and vital signs from electronic health records of adult patients over four years (n = 133,089 encounters). We analyzed primary outcomes of unplanned intensive care unit transfer, emergency vasoactive medication administration, cardiac arrest, and death. We compared PICTURE with existing early warning systems and logistic regression at multiple levels of granularity. When analyzing PICTURE on the testing set using all observations within a hospital encounter (event rate = 3.4%), PICTURE had an area under the receiver operating characteristic curve (AUROC) of 0.83 and an adjusted (event rate = 4%) area under the precision-recall curve (AUPR) of 0.27, while the next best tested method—regularized logistic regression—had an AUROC of 0.80 and an adjusted AUPR of 0.22. To ensure system interpretability, we applied a state-of-the-art prediction explainer that provided a ranked list of features contributing most to the prediction. Though it is currently difficult to compare machine learning–based early warning systems, a rudimentary comparison with published scores demonstrated that PICTURE is on par with state-of-the-art machine learning systems. To facilitate more robust comparisons and development of early warning systems in the future, we have released our variational autoencoder’s code and weights so researchers can (a) test their models on data similar to our institution and (b) make their own synthetic datasets.

**Highlights:**

- Novel simulation shows that learning missingness patterns in EHR data decreases early warning system performance if missingness pattern changes
- Simulation generated synthetic EHR data using variational autoencoder with custom loss function to account for high missing rate
- Randomized imputation and Bayesian regression imputation prevented tree-based methods from learning missingness patterns
- Using appropriate imputation, we developed PICTURE, an early warning system for patient deterioration
- PICTURE performance is comparable to currently used systems and it can explain predictions via feature ranking

**Graphical Abstract:** 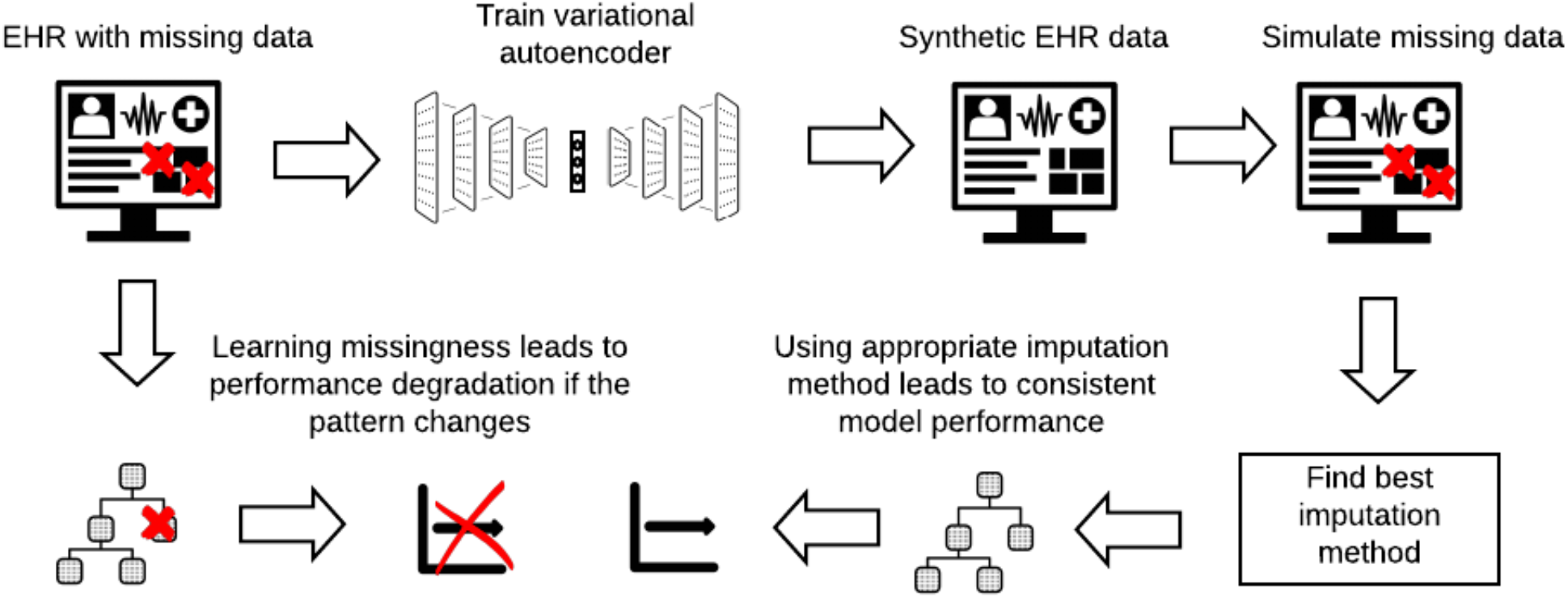

## 1. INTRODUCTION

Of all unplanned patient transfers to intensive care units (ICUs), approximately 90% of these transfers may occur due to a new or worsening condition [1], [2]. However, it is likely that around 44% of patients’ adverse events in the hospital are avoidable [3]. Moreover, an estimated 85% of severe adverse events are preceded by abnormal physiological signs [4], which may present 24–48 hours in advance [5]. This means we can develop early warning systems (i.e., predictive data analytics) that leverage the information in those abnormal signals to alert providers of impending patient deterioration, providing an opportunity for preventive treatment or escalation of care to the ICU. Early warning systems are an important tool for secondary prevention (e.g. early recognition of sepsis) and tertiary prevention (e.g. informing in-hospital critical care to minimize the morbidity) [6]. Thus, an early warning system that analyzes individual patient records is an important component of digital preventive healthcare because it would ideally reduce critical care needs. If successful, early warning systems may ultimately lead to improved care and reduced costs [7], [8].

Despite the importance of these early warning systems, it is currently difficult to compare these tools between different medical institutions, and even more importantly, they do not always generalize well beyond the hospital systems they were developed in. This is especially true for machine learning-based systems, which have increased predictive power over commonly used expert-defined aggregate scores [1] and linear methods like logistic regression for predicting deterioration and sepsis [9]–[15]. Specifically, when systems are trained at one institution, their performance may drop when they are applied to another institution’s data [1]. This may occur for two reasons. First, the machine learning system might be learning the institution-specific patient population characteristics and ICU transfer policy. Thus, if the patient population and the ICU transfer policy differs at the other institution, the performance could drop. The second potential cause has not been sufficiently explored in the literature: the machine learning system may be learning missingness patterns in the data. Electronic health record (EHR) data has many characteristics (e.g. collection of data for certain treatments) that may bias early warning system development [16] and the missingness pattern may capture these characteristics. Specifically, tree-based machine learning methods have the potential to learn the missingness pattern when using methods such as mean or extreme value imputation. Some have suggested that learning missingness patterns improves performance [17], which is true when the pattern is correlated with the target (e.g. adverse events) and when the pattern remains the same across datasets. However, as we demonstrate here, performance actually worsens when the missingness pattern changes (e.g., when the model is used in a different hospital setting or policy changes cause different missingness patterns). Using an insufficient imputation strategy during model development may therefore lead to false confidence in the general performance of a tree-based early warning system, and it may prevent the early warning system from generalizing to other medical systems. Furthermore, certain missingness patterns may not provide new information to care providers. For example, missingness patterns in the presence or absence of certain laboratory tests are not useful because providers already know if they have ordered a test or not. If an early warning system alerts based on this specific missingness pattern, it is possible that an alert would go out simply because a certain test is ordered. This would not be useful and would contribute to alarm fatigue, providing another compelling reason to ensure early warning systems are not learning these missingness patterns. To ensure that predictive analytics are appropriately developed for preventative healthcare, it is important that researchers understand this problem and use appropriate imputation methods.

As presented in this paper, we developed a novel simulation to demonstrate the existence of the learning missingness problem and its effects on generalization performance. Recently, there has been an upsurge in interest in synthetic patient data and anonymized machine learning of patient data [18]–[22]. However, these methods have not previously been used for the specific context of early warning system issues of learning missingness patterns. We thus uniquely leveraged synthetic patient data to develop a simulation and discover a solution to the problem of learning missingness. Our novel simulation used a variational autoencoder [23] with a novel loss function to handle a high amount of missing data, which is characteristic of EHR data. Using this simulation, we identified imputation methods that prevent tree-based early warning systems from learning missingness patterns. Then, using an appropriate imputation method, we developed a new early warning system (PICTURE: **P**redicting **I**ntensive **C**are **T**ransfers and other **U**nfo**R**eseen **E**vents) to predict patient deterioration in the hospital, which will aid in tertiary preventive healthcare.

## 2. Materials and Methods

### 2.1. Setting and Study Population

We used four years (2015–2018) of EHR data from a large academic tertiary referral system (Michigan Medicine). We used the data from 2015–2017 to train the autoencoder and simulate EHR data, which allowed us to identify the imputation methods that do not learn missingness patterns. We then used all four years of data (2015–2018) to train, tune hyperparameters, and test for our early warning system. **Table 1** describes the study cohort and event rates. Inclusion criteria included adult patients (age ≥18) classified as inpatient, observation, outpatient surgery, or outpatient in a bed. For inclusion, we required a complete blood count, basic metabolic panel, and at least one vital signs order. We excluded patients with a ventricular assist device. The study protocol was approved by the University’s Institutional Review Board.

**Table 1:** Cohort Summary.

| Demographics | Training and Validation Cohorts (n = 90,185) | Testing Cohort (n = 42,987) | Total (n = 133,089) |
| --- | --- | --- | --- |
| **Years** | 2015-2017 | 2018 | 2015-2018 |
| **Age, mean (SD)** | 57 (18) | 57 (18) | 57 (18) |
| **Female sex, n (%)** | 44,713 (50%) | 21,604 (50%) | 66,317 (50%) |
| **Race** |  |  |  |
| Asian, n (%) | 1,849 (2%) | 848 (2%) | 2,697 (2%) |
| Black, n (%) | 9,803 (11%) | 5,303 (12%) | 15,106 (11%) |
| Other, n (%) | 3,892 (4%) | 1,970 (5%) | 5,862 (4%) |
| White, n (%) | 74,558 (83%) | 34,866 (81%) | 109,424 (82%) |
| **ICU transfers, n (%)** | 2,692 (3%) | 1,043 (2%) | 3,735 (3%) |
| **Cardiac arrests, n (%)** | 207 (0%) | 55 (0%) | 262 (0%) |
| **Vasoactive meds, n (%)** | 1,947 (2%) | 520 (1%) | 2,467 (2%) |
| **Death, n (%)** | 1,473 (2%) | 432 (1%) | 1,905 (1%) |

### 2.2. Predictors

Based on the predictors included in prior early warning systems and the expertise of the clinicians on our team, we used the following predictors collected from the EHR (EPIC, Verona, WI) for our simulation and PICTURE model development: vital signs (hemoglobin oxygen saturation, heart rate, systolic and diastolic blood pressure, respiratory rate), oxygen supplementation status, Glasgow coma score, urine output over 24 hours, complete blood count, basic metabolic panel, less common labs (bilirubin, partial thromboplastin time, prothrombin time, international normalized ratio, lactate, magnesium, phosphorous, albumin, protein-level), and calculated values (shock index, mean arterial pressure, age multiplied by shock index, pulse pressure). Fluid bolus administration was also included as a binary indicator variable. We also included race by one-hot encoding it (where we have indicators for White, Black, Asian, or Other), a sex indicator variable (Female), and age at the start of the encounter. We coded race this way based on the frequency of the categories, and all races with a frequency < 1% were coded as “Other.” **Table D.1** shows the full list of predictors, the mean, the standard deviation, the overall missing rate, and the missing rate stratified by adverse events.

### 2.3. Outcomes

Primary outcomes for the simulation and PICTURE model were ICU transfer from a general ward or telemetry unit, death, cardiac arrest (following the American Heart Association’s Get With The Guidelines^®^), and vasoactive medication administration (i.e., epinephrine, norepinephrine, dopamine, dobutamine, vasopressin, milrinone, phenylephrine). We determined ICU transfer by inspecting location information and patient accommodation level. We considered the general ward to be any floor that admitted patients and was not the emergency department, the operating room, nor the intensive care unit. We annotated vasoactive medication initiation events using medication administration orders, and clinicians manually identified order codes associated with emergency vasoactive medication administration. Vasoactive administration is an important outcome because it can precede an ICU transfer. By including vasoactive medication administration, our model was able to learn deterioration events earlier, before they were indicated by ICU transfer. Cardiac arrests were also a useful deterioration target (over and above death) since not all cardiac arrests result in death. Throughout the paper, we refer to the set of all outcomes as the “outcome” or “target.”

### 2.4. Population Refinement for Unplanned ICU Transfers

The purpose of our PICTURE early warning system is to identify unplanned ICU transfers to aid in tertiary preventive healthcare. However, we did not want our model to predict *planned* ICU transfers, which are common. Thus, we excluded patients who were directly admitted to an ICU (e.g. admitted from the emergency department or from another hospital). We also excluded patients who were directly transferred to an ICU from an operating room, a catheterization laboratory, or interventional radiology unit. Only the first general ward stay per hospital encounter was used.

### 2.5. Model Development

To partition our data for model development, we followed the structure from Brian Ripley [24]. In Ripley’s scheme there is a training set to learn model parameters, a validation set to tune hyperparameters, and a test set to estimate model performance. This three set split is common for state-of-the-art deep learning models [25]. Also, the objective of an early warning system is to predict the future, so a time-based partitioning of patient hospital encounters fits this framework. We thus split our data by time as follows: encounters from 2015–2017 were used for training and validation/hyperparameter tuning (n = 90,185; 68% of total encounters in our dataset). Specifically, 80% of patients from 2015–2017 were used for training, and 20% were reserved for validation and hyperparameter tuning. All encounters from 2018 were used for testing and estimating model performance (n = 42,987; 32% of total encounters in our dataset) only after the models were finalized (**Figure D.1**). If patients in the 2018 test set had prior hospital encounters in 2015–2017, we discarded those encounters in the 2015–2017 training and validation sets to reduce bias by a patient being in both the training and test set. We excluded all observations 30 minutes before the first adverse event or later to be consistent with other published approaches [26]. Based on our simulation results described below, we imputed the data set for training PICTURE by using a Bayesian regression and taking the mean of the posterior. XGBoost[27], a gradient boosting tree, was used as the classification model in PICTURE and analyzed in the simulation. Additional details related to training the PICTURE analytic are in **Appendix B.1**.

### 2.6. PICTURE Model Evaluation

#### 2.6.1. Comparing to Other Early Warning Systems

Using our test set, we compared PICTURE to two expert-defined aggregate scores via within-study comparisons: the National Early Warning Score (NEWS) [28], [29] and the Sequential Organ Failure Assessment Score (SOFA) [30]. We chose NEWS and SOFA because these are commonly used and easy to compute [1], [31]. We implemented NEWS and SOFA scores using their published scoring systems. We also compared PICTURE to a logistic regression model using the same features and imputation method. For the logistic regression model, we used a regularized model (L2) where we selected the optimal penalty term on the validation set. To compare PICTURE to other published machine learning–based early warning systems for patient deterioration, we had to take a different approach because most do not provide source code or data. Thus, we compared PICTURE’s performance to published results for machine learning–based early warning systems using standard criteria to show our model has comparable performance. We compared PICTURE to the following models: one by Alvarez et al. [13], the electronic cardiac arrest risk triage (eCART) [26], eCART random forest [9], [14], and the Advanced Alert Monitor [10], which were highlighted in recent literature reviews [1],[16]. Because we could only compare our performance indicators to published performance indicators, this does not allow us to definitively determine which score performs the best.

#### 2.6.2. Granularity Levels

We used three granularity levels in this study. For training, we used the window level, which decomposed a patient’s encounter into a series of 8-hour blocks. During testing, we compared PICTURE to a logistic regression model, NEWS, and SOFA at two other levels of granularity: the observation level and the encounter level (**Figure D.2**). The observation level corresponds to every time relevant data was collected from a patient. Observations were labeled ‘1’ if they occurred up to 24 hours before the event and labeled ‘0’ otherwise. The encounter level corresponds to making a single prediction for a patient’s entire hospital encounter. We examined the encounter level predictions by taking the maximum score across each encounter up to the first adverse event if one occurred (“maximum”). For both encounter-level assessments, all observations after the first adverse event were removed from the dataset.

It is important to note that the three levels of granularity (window, observation, and encounter) all have different event rates. The event rate for the window-level event rate corresponds to the fraction of windows with an event. The observation-level event rate corresponds to the fraction of observations up to 24 hours before an event. The encounter-level event rate corresponds to the fraction of encounters with an event. Most commonly, the encounter-level event rate is reported. These event rates are important for any performance measures related to the precision of the classification model.

#### 2.6.3. Performance Indicators

We used the area under the receiver operating characteristic curve (AUROC) and the area under the precision-recall curve (AUPR) as the primary criteria to compare PICTURE with NEWS and SOFA. The AUPR quantifies the positive predictive value (PPV) averaged across all sensitivities. To add confidence intervals around the AUPR and AUROC, we used a bootstrap method. Specifically, we used 500 replications to compute pivotal confidence intervals 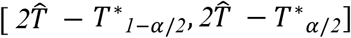, where 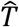 was the statistic, *T*\*_*x*_ was the resampled test statistic at quantile *x*, and *α* was the confidence level set at 0.05. For observation-level results we used a block-bootstrap. The event rate was standardized to 4% for comparability with published literature using the formula: 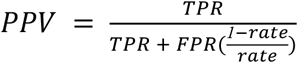. To compare PICTURE to other machine learning systems, we could not use AUPR because only one other system reported it [10], so we inspected the PPV and the workup-to-detection ratio (WDR), which indicates the number of alarms that will sound for every true deterioration event (meaning lower WDR is better). PPV and WDR were analyzed at 51% sensitivity for comparison purposes. It is important to note that while WDR is chosen for a specific sensitivity level, AUPR is the average PPV across all sensitivity levels. So WDR is useful for summarizing the performance at a specific threshold, while AUPR is useful for summarizing a model’s performance across many thresholds.

### 2.7. Simulation Design

Prior to developing and evaluating the PICTURE model as described above, we built a variational autoencoder that generated synthetic data that looked similar to our training dataset. This allowed us to show how tree-based classifiers can learn missingness patterns and how to eliminate the problem. We then assessed the effects of different imputation methods on a variety of missing data settings (described below) to understand the extent of the problem and the degree to which imputation methods ameliorated the problem.

#### 2.7.1. Simulation Input Data Formatting

To create a model that sufficiently showed the extent of the missingness problem, we analyzed data at the encounter level. To select a single set of predictors per encounter, we started with the windowed training dataset. We then split the dataset into encounters with an adverse outcome and those without an adverse outcome. For encounters with an adverse event, we took the window that was labeled ‘1,’ and for encounters without an adverse event we selected the window with the highest NEWS score. In other words, we selected windows where the first adverse event occurred, or if one did not occur we selected windows where the patient was the sickest as defined by NEWS. This resulted in one training example per encounter, and these nonevent encounters (when NEWS is highest) are the most challenging windows to classify per encounter because they are the most similar to the adverse event windows. We included all the same variables as listed above with the exception of race and gender.

For the training data, we scaled the data to have a mean of zero and a standard deviation of one. Finally, we imputed data using the mean of each variable, which was zero. We chose mean imputation for the input of the simulation to ensure we did not bias the simulation results toward a different imputation method.

#### 2.7.2. Variational Autoencoder For Synthetic Data Generation

Variational autoencoders are a variant of deep-stacked autoencoders. Autoencoders are a type of deep learning model that tries to learn a compact representation of their input data. It is essentially a deep neural network with the same inputs and outputs. The primary benefit of these models is that they can learn a compact non-linear representation of the input data. Autoencoders have an encoder that takes the input vector and computes the coding, and they have a decoder that takes the coding and tries to regenerate the input. Variational autoencoders extend these models by making them generative models. This occurs by making the encoder learn the mean and variance of a distribution that models the input vector. During training one can sample randomly from the distribution using the mean and variance output from the encoder and compute the loss function using the reconstruction computed from this random sample. Finally, the loss function has an extra term that calculates the Kullback-Leibler distance from the means and variances of the codings to a spherical Gaussian distribution with a mean of zero and a variance of one. To generate novel data, one simply has to sample from a *c* dimensional spherical Gaussian distribution with a variance of one, where *c* is the dimension of the coding vector, and apply the decoder.

There were two things we needed to customize for this model to work well for EHR data, given its high missing rate and our desire to simulate both continuous predictors and the binary target variable. First, we needed to specify a loss function to work with continuous and binary variables. Second, due to the high missingness inherent to EHR data, we excluded imputed values in the loss function reconstructions because this would bias the results. **Appendix B.2** shows the complete formulation and justification for our variational autoencoder’s custom loss function.

We followed a symmetrical structure, which is common for autoencoders [32]. To add additional randomness to the input, we applied dropout at 15% for input predictors. **Figure 1** shows the structure of the autoencoder. We tied the weights of the network layers to make the network symmetrical and reduce the number of parameters. The coding size (*c*) was 16, and we used an ‘Adam’ optimizer to fit the model with early stopping, with a ‘patience’ of 7 epochs, a batch size of 512, and a maximum of 100 epochs. The final inspection of reconstruction performance was done by comparing the observed and reconstructed feature correlation matrices. We fit the model using *keras* (version 2.3.1) with a *tensorflow* (version 2.0.0) backend.

**Figure 1:**
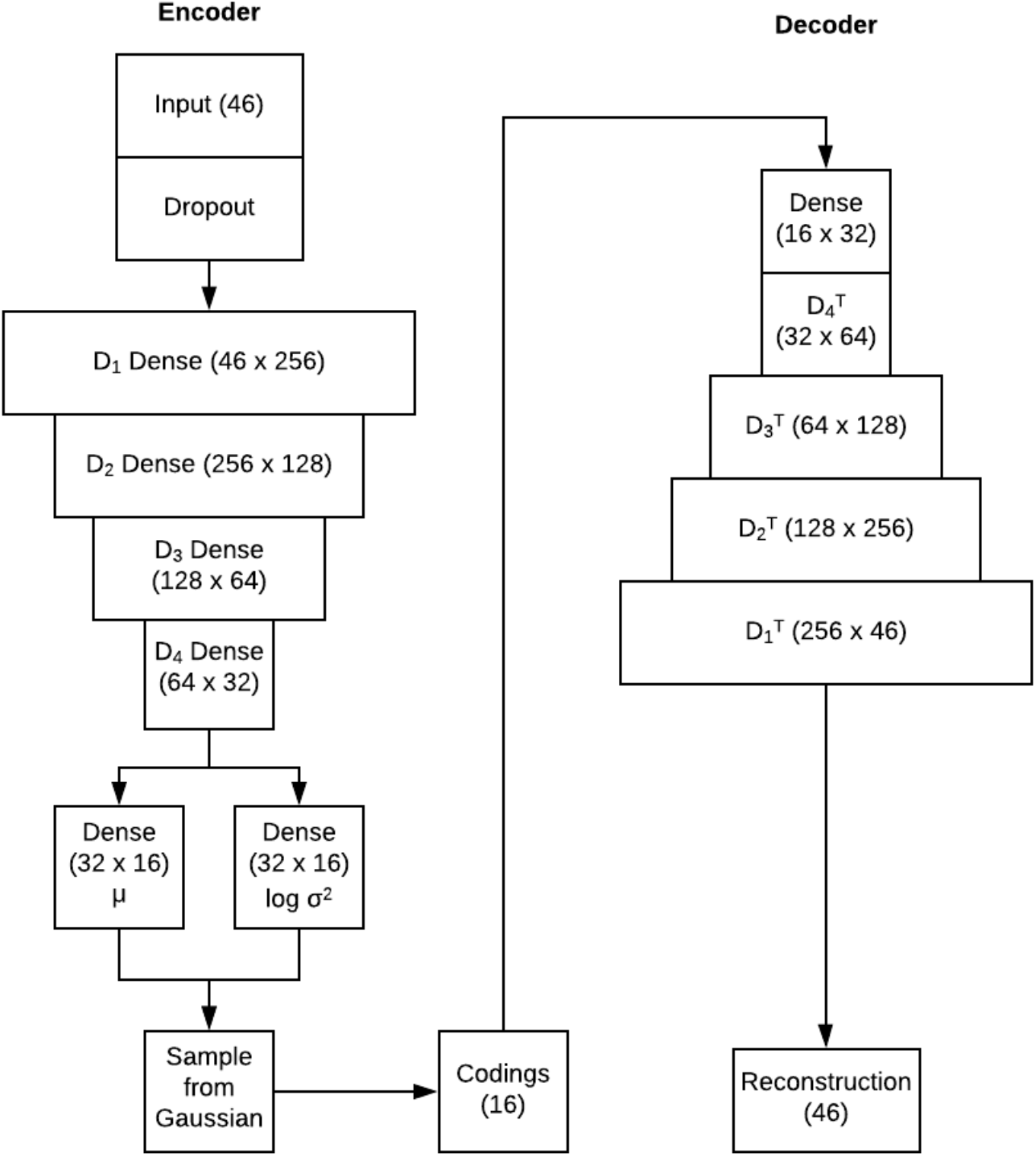
Variational Autoencoder Structure. The left side has the encoder and the right side has the decoder. For the dense layers, the numbers in parentheses correspond to the shape of the weight matrices, while the number in the parentheses for the input, codings, and reconstruction boxes correspond to the shape of the vector. In the encoder, an input vector with 45 features had input features randomly dropped out at 15%. The result of this was then sent through four dense layers, which ultimately computed two vectors of size 16: one corresponding to the mean and the other corresponding to the log variance of the distribution of coding corresponding to the input vector. To get the coding for the input, we randomly sampled from the distribution. This coding was passed into the decoder, which has a dense layer that maps the coding size to the appropriate size. This was then passed into the transposed layers from the encoder, which resulted in a reconstruction the same shape as the input vector. In the dense layers, we used the Scaled Exponential Linear Unit [33] activation function, and for the binary target output we used the sigmoid activation function.

#### 2.7.3. Simulation for Comparing Imputation Methods for Missingness Pattern Changes

Our aim of the simulation was to show how learning missingness patterns will lead to a performance drop when the missingness pattern changes, and how to avoid this issue. To do this, we created four scenarios where we trained models under one missingness situation and applied the model under different missingness patterns. In Scenario 1, the training data and the test dataset followed the same missingness pattern of the observed data. **Table D.1** shows the missingness pattern per class. These are the values used for Scenario 1. In Scenario 2, there was half as much missing data in the testing dataset as in the training dataset. In Scenario 3, the missingness pattern in the testing dataset was fixed across classes (i.e., those with and without adverse events), but it was different in the training dataset. This situation represents an example where a variable such as lactate is taken more frequently in a different setting and is no longer correlated with the severity of illness. In Scenario 4, the testing dataset was fully observed.

We evaluated the following classification models: XGBoost, random forest, logistic regression, and logistic regression fit using missing indicators only. The missingness patterns were constructed using our observed training dataset from Michigan Medicine. Variables were set to be missing using a simple pattern. First, we computed the missing rate per variable in encounters with an event and those without. Second, we altered the missing rate per class according to the scenarios above. Third, after we generated data from the variational autoencoder, we set observations to be missing according to the missing rate per class. This model assumes the variables that are set to be missing are conditionally independent given the class label (adverse event status).

We evaluated the following imputation methods: 1) “None” corresponds to just leaving the missing values (used for XGBoost only); 2) “Extreme Value” corresponds to imputing −100; 3) “Mean” corresponds to imputing the mean; 4) “Random Healthy” corresponds to imputing data from the healthy patients’ empirical distribution limited to 1-percentile and 99-percentile; 5) “Bayesian Regression Random” corresponds to fitting a Bayesian regression model using observed variables and sampling randomly from the posterior to estimate the missing values; and 6) “Bayesian Regression Mean” is the same except the mean of the posterior is used. **Figure 2** shows the overall structure of the simulation comparison.

**Figure 2:**
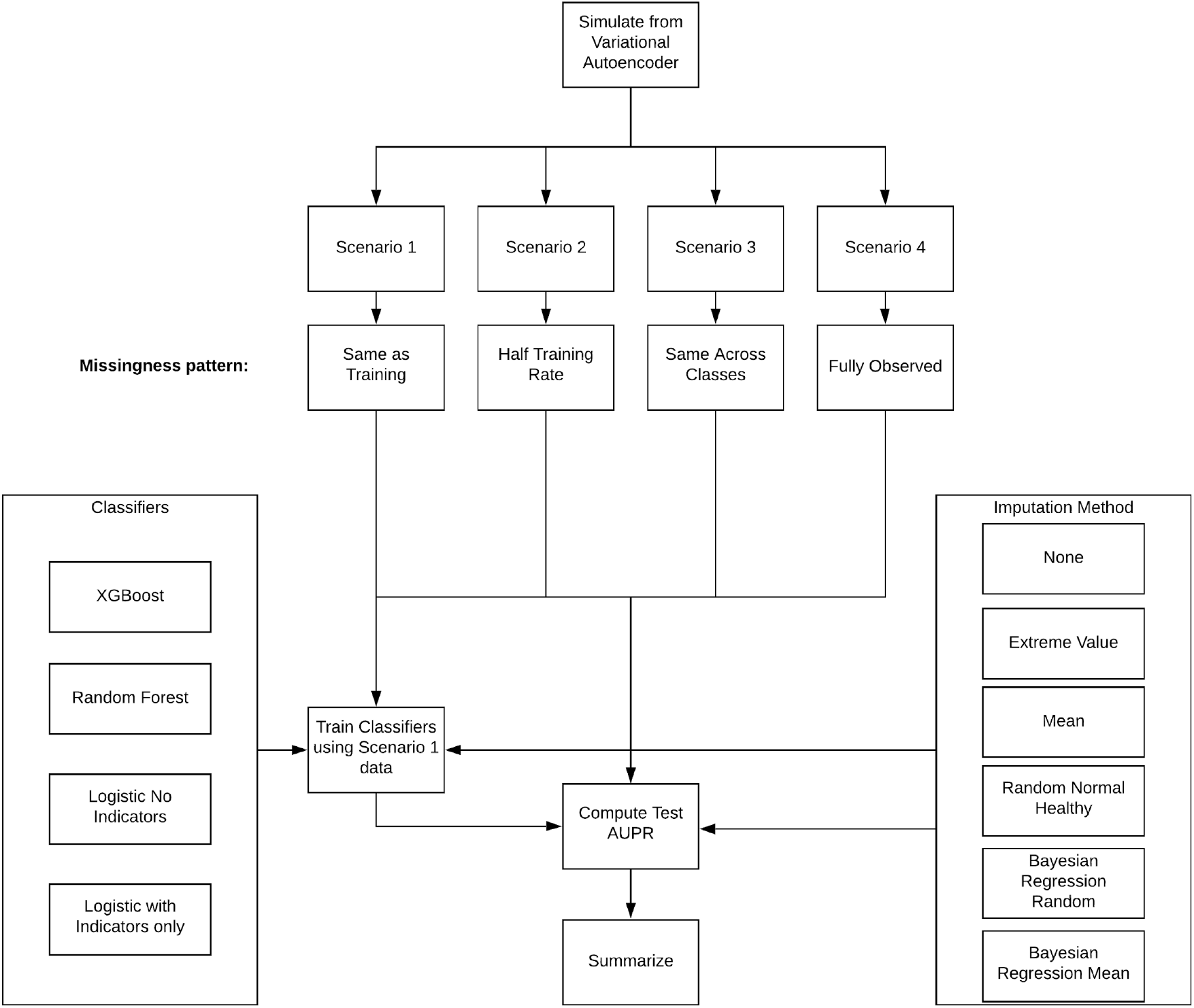
Simulation Design. We generated simulated training, validation, and testing datasets from the variational autoencoder. We applied four missingness patterns that represent different scenarios. We trained all classifiers with all the imputation methods (where applicable), using the Scenario 1 training dataset and applying the classifiers to the testing datasets from each Scenario.

### 3. Results and Discussion

#### 3.1. Simulation Results

All the results presented in this section refer to the test set performance unless otherwise noted. For the simulation study, the simulated testing set consisted of the same patients across scenarios with different missingness patterns.

##### 3.1.1. Our Variational Autoencoder Produces Data Similar to Michigan Medicine’s Data

A precondition to assessing the simulation results was to verify that the simulation model produced realistic samples. To assess this, we compared the raw Michigan Medicine validation data with the reconstructions produced by our variational autoencoder. We first compared the pairwise Pearson correlation matrix between variables in the raw data and we did the same for the variational autoencoder reconstructions. We found that 88% of correlations from the upper triangle of these matrices were in the same direction. Second, we computed the Pearson correlation between the raw data and reconstructions for each variable. We found the average Pearson correlation across all variables was 0.76. These results suggest that the variational autoencoder produced data similar to the real Michigan Medicine data and that it is a reasonable simulation model.

To assess the improvement from our custom loss function over the standard mean square error loss, we performed the same average Pearson correlation analysis. Using the mean square error loss, we obtained an average Pearson correlation of 0.69 as compared to 0.76 for our custom loss function. For variables with a missing rate > 50%, the average Pearson correlation dropped to 0.62 for the mean square error loss function. When using our custom loss function, the average Pearson correlation for these variables was 0.81. To further elucidate these results, we looked at the Pearson correlation between the raw data and reconstructions for lactate. When using the mean square error loss, the Pearson correlation was 0.56. When using our custom loss function, the Pearson correlation for lactate was 0.8. Therefore, our custom loss function improved the learning across variables with a higher missing rate, making it applicable for EHR data.

##### 3.1.2. Randomized and Bayesian Regression Imputation Prevents Tree-Based Models from Learning Missingness Patterns

The purpose of this analysis was to show that: 1) tree-based methods can learn missingness patterns in EHR data; 2) learning missingness patterns is undesirable if the pattern may change; and 3) to determine which methods mitigate the problem. Learning the missingness pattern can improve performance if the pattern is correlated with the target or outcomes of interest (**Table D.1**). Missingness patterns are correlated with patient deterioration in EHR data because clinicians order less common labs (e.g., lactate) when they are concerned about a patient. The difference in missing rate between patients with the adverse event and without corresponds to the strength of the correlation between the missingness pattern and the adverse event. Early warning systems that exclude variables with a high missing rate will be less affected by this issue.

We found that Bayesian imputation strategies and randomized imputation strategies address the issue. **Figure 3** shows the results comparing the AUPR of different classification models and imputation strategies across the four scenarios. When using the “Extreme value” imputation method, XGBoost’s test set AUPR dropped from 0.86 in Scenario 1 to 0.15 in Scenario 3. The Random Forest classifier’s performance dropped from 0.61 in Scenario 1 to 0.19 in Scenario 3 using the same imputation method. The pattern was the same for the “None” and “Mean” imputation methods. Logistic regression did not see this same performance drop for “Mean” imputation. For illustrative purposes, we show the performance for “Extreme Value” imputation for logistic regression as it was being used as an indicator, and thus the AUPR dropped as the missingness pattern changed from 0.81 in Scenario 1 to 0.07 in Scenario 3. Again, if we only used indicator variables in a logistic regression model, then a similar performance drop was observed. Therefore, tree-based classifiers can learn the missingness pattern in the data, and logistic regression can learn the missingness pattern through indicator variables. Learning these patterns leads to increased performance if the missingness pattern is consistent; however, the performance drops if the missingness pattern changes.

**Figure 3:**
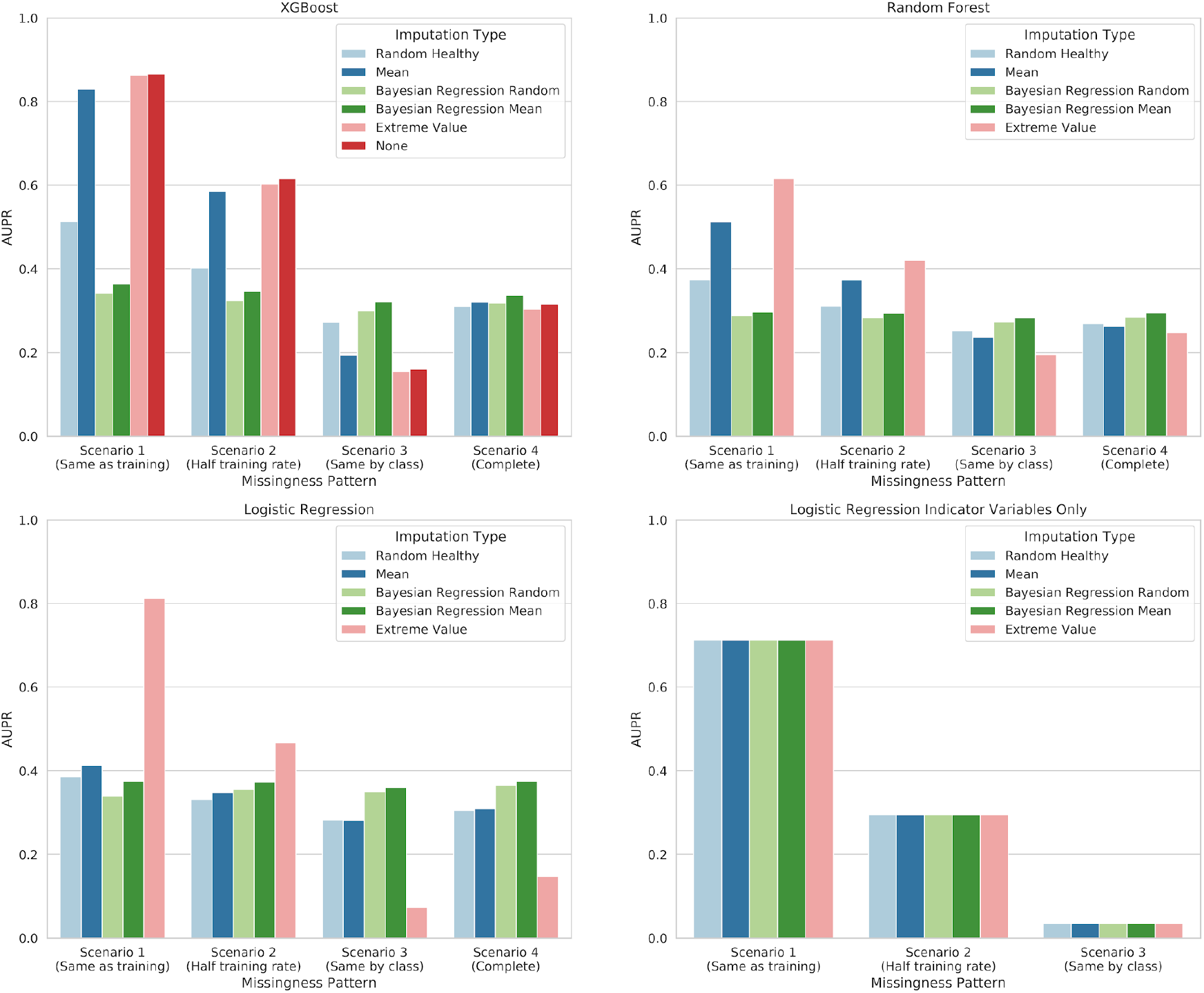
Simulation results comparing test performance of XGBoost, a Random forest classifier, logistic regression, and logistic regression with missing value indicators as features across different test setting missingness patterns. The same imputation method that was used in training was also used during testing. In Scenario 1, the training data and the test dataset followed the same missingness pattern: the observed data. In Scenario 2, there was half as much missing data in the testing dataset as in the training dataset. In Scenario 3, the missingness pattern in the testing dataset was fixed across classes, and it was different in the training dataset. In Scenario 4, the testing dataset was fully observed. These different missing patterns could represent different hospitals or data collection changes. Each panel represents a different classification model and each color within a panel represents a different imputation method. The learning of the missingness pattern is most clearly illustrated in the XGBoost classification model with the “None” imputation method, where the AUPR was 0.87 in Scenario 1 and 0.16 in Scenario 3. Note that these are the same simulated test patients with a different missingness pattern. Bayesian imputation strategies eliminated the learning of the missingness pattern.

The Bayesian imputation methods consistently had the highest performance across simulated scenarios, except the indicator only model, which is just using indicators and not imputed values so it is invariant to the imputation method. “Random Healthy” mostly eliminated the learning of the missingness pattern, but its performance was lower than the Bayesian methods. Because the “Bayesian Regression Mean” method performed the best in our simulation and eliminated the missingness pattern learning for tree-based models, we used this imputation method for PICTURE.

We then investigated the sensitivity of the tree-based classifiers to learning the missingness patterns using different missing rates. In **Figure D.3**, we trained the models using data with half the missing rates of those listed in **Table D.1**. The tree-based classifiers still learned the missingness pattern and experienced a performance drop when the missing rate decreased and a performance increase when the missing rate increased. This is contrary to what we typically expect, because usually performance worsens as the missing rate increases. This didn’t happen here because the missingness pattern is what the models learned, so when the missingness increased the performance improved. In **Figure D.4**, we trained the models with four times lower missing rates than those listed in **Table D.1**. A less pronounced drop in performance occurred, but interestingly the Random Forest model was only marginally affected by learning the missingness pattern regardless of the imputation method.

It seems plausible that the use of the Bayesian imputation methods will improve performance across all models, including expert-defined scores like NEWS, since they are estimating missing values closer to the truth as compared to “Mean” imputation. This is evidenced by how the performance was the highest in Scenario 4 (the fully observed case) when training using the “Bayesian Regression Mean” method. This suggests that the training cohort, when using this imputation approach, was most similar to the fully observed test cohort, which is why the classification algorithms performed the best in this scenario. In **Appendix C.1**, we also show a simpler scenario where tree-based models can learn missingness patterns even with a single predictor. The primary result is that the larger the difference in missing rate between classes (i.e., those with vs. those without adverse events), the larger the performance decrease when the missingness pattern changes.

##### 3.1.3. Learning Missingness Patterns is Not Always Desirable

Some have suggested that learning missingness patterns is useful for predictions [17]. We agree with that sentiment; however, it is only useful in certain situations. If the missingness pattern is not expected to change, then including missing value indicators is useful and will improve predictive performance if the indicator is correlated with the class label. However, if the missingness pattern changes, our simulation shows that performance will drop. This problem is important for early warning system developers to know about and address, provided they expect the missingness pattern to change. Another reason that learning missingness patterns is undesirable for early warning systems is that clinicians order tests because they believe a patient is sick, so if a model’s predictions are driven by the presence or absence of a lab, it is not providing novel information to the clinician (see an example of this in **Appendix C.1**). It is especially important for researchers to address this when developing new predictive analytics at a single center with the intention to generalize to other centers, because it is plausible that missingness patterns will be different across institutions.

##### 3.1.4. The Variational Autoencoder and Simulation have Several Benefits

Our variational autoencoder with its novel loss function and our simulation were useful tools for generating data similar to our observed Michigan Medicine cohort. Because these tools could be used to generate synthetic data similar to other institutions’ actual data as well, this could be useful for comparing the performance of algorithms trained at different institutions. For example, if other institutions used the simulation and variational autoencoder to produce their own synthetic data, they could then release their synthetic data, and the collection of synthetic data from a variety of institutions would allow various proprietary machine learning–based early warning systems to be more robustly compared on the same data. This would simultaneously advance the field more efficiently while alleviating health record privacy concerns. Other groups are developing similar ideas for sharing EHR data [34]. To this end, we have released our variational autoencoder’s code, weights, and the synthetic data we generated (https://github.com/MCIRCC/picture-simulation).

In addition to allowing more robust comparisons of various early warning systems, releasing our code will also provide several other benefits. For example, it has been shown that combining synthetic data with real data improves test performance [35], [36]. Therefore, our variational autoencoder could further improve other researchers’ predictive analytics for preventive healthcare by allowing them to train on synthetic data, and they could make their models more robust by adding missingness to the synthetic data. Furthermore, the simulation could be useful for comparing new imputation methods under diverse missingness patterns. While this will be very useful, it is important to note that the synthetic data from the variational autoencoder is mathematically modeled and thus easier to classify than actual noisy data.

#### 3.2. PICTURE Performance Results

All the results presented in this section are from our 2018 testing cohort. This cohort was not used in training the model, and it was not used for hyperparameter selection. There is no patient overlap as determined by medical record number between the training/validation cohort and the testing cohort. Beyond the results presented here, we show additional analyses that further illustrate the clinical impact with regards to how early PICTURE makes a prediction, how well it performs in subgroups and feature interaction explanations in **Appendix C.2**.

##### 3.2.1. PICTURE Performs Better than NEWS, SOFA, and Logistic Regression in a Michigan Medicine Cohort

**Table 2** shows comparisons between PICTURE, logistic regression, NEWS, and SOFA for predicting general deterioration at the observation and encounter levels. We also calculated the differences in AUROC and AUPR using the raw event rate at the observation level. The PICTURE AUROC was significantly higher than the logistic regression AUROC by 0.032 [CI: 0.025, 0.039], higher than the NEWS AUROC by 0.063 [CI: 0.052, 0.072], and higher than the SOFA AUROC by 0.204 [CI: 0.164, 0.233]. The PICTURE AUPR was significantly higher than the logistic regression AUPR by 0.028 [CI: 0.023, 0.035], higher than the NEWS AUPR by 0.022 [CI: 0.012, 0.034], and higher than the SOFA AUPR by 0.068 [CI: 0.057, 0.079] with an observation-level event rate of 1% ( **Figures D.5-D.7**). Using encounter-level granularity and setting the sensitivity at 51% with a raw event rate of 3.4%, the WDR for PICTURE was 3.9, meaning a true alarm would occur once for every four alarms, while logistic regression had a WDR of 5.3, NEWS had a WDR of 6.1, and SOFA had a WDR of 11.3. To verify PICTURE performance, a group of clinicians and data scientists manually verified a subset of the results.

**Table 2:** PICTURE, Logistic Regression, NEWS, and SOFA performance comparison at the observation level and encounter level.

| Method | N (rate %) | AUROC | Raw AUPR | AUPR (rate=4%) |
| --- | --- | --- | --- | --- |
| Observation Level |  |  |  |  |
| PICTURE | 4,581,561 (1%) | 0.833 [0.824, 0.841] | 0.098 [0.088, 0.108] | 0.271 [0.250, 0.290] |
| Logistic Regression | 4,581,561 (1%) | 0.801 [0.787, 0.816] | 0.071 [0.063, 0.078] | 0.218 [0.200, 0.233] |
| NEWS | 4,581,561 (1%) | 0.770 [0.753, 0.784] | 0.077 [0.062, 0.088] | 0.217 [0.189, 0.241] |
| SOFA | 4,581,561 (1%) | 0.631 [0.607, 0.664] | 0.031 [0.025, 0.036] | 0.103 [0.089, 0.117] |

| Encounter Level (maximum) |  |  |  |  |
| --- | --- | --- | --- | --- |
| PICTURE | 42,987 (3.4%) | 0.856 [0.844, 0.866] | 0.286 [0.263, 0.308] | 0.314 [0.290, 0.336] |
| Logistic Regression | 42,987 (3.4%) | 0.812 [0.800, 0.824] | 0.195 [0.178, 0.211] | 0.219 [0.199, 0.234] |
| NEWS | 42,987 (3.4%) | 0.777 [0.763, 0.793] | 0.213 [0.191, 0.234] | 0.235 [0.214, 0.258] |
| SOFA | 42,987 (3.4%) | 0.686 [0.671, 0.701] | 0.141 [0.125, 0.158] | 0.157 [0.140, 0.174] |
| In addition to providing the AUPR using the raw adverse event rate, we provide the AUPR at a 4% event rate to standardize for cross-study comparison. |  |  |  |  |

##### 3.2.2. PICTURE Has Comparable Performance to Other Published Machine Learning Systems

One major challenge for comparing PICTURE to published machine learning–based methods is that the code is not available for the models. Also, the datasets that other models were tested on are not available due to privacy concerns related to EHR data. This makes it challenging to show that one method is better than another, so our goal was simply to compare performance metrics. **Table 3** shows that PICTURE performance was comparable to the published performance metrics of other state-of-the-art machine learning–based systems.

**Table 3:** Performance comparison between PICTURE and other published machine learning systems.

| Comparison | Alvarez et. al [13] | eCART [26] | eCART random forest [9], [14] | Advanced Alert Monitor [10] | Average of published systems | PICTURE |
| --- | --- | --- | --- | --- | --- | --- |
| Number of Test set Encounters | 3,792 | 107,911 | 107,911 | ~162,350 |  | 42,987 |
| Classifier Type | Logistic regression | Logistic regression | Random Forest | Logistic regression |  | Gradient Boosting Tree |
| Granularity Level | 24-hr window (patient days) | Observation | Observation | Encounter |  | Observation |
| AUROC (95% CI) | 0.85 | 0.77 | 0.8 | 0.82 | 0.81 | 0.83 |

|  | (0.82-0.87) | (0.76-0.77) | (0.80-0.80) | (0.81-0.83) |  | (0.82-0.84) |
| --- | --- | --- | --- | --- | --- | --- |
| **Sensitivity** | 0.52 | 0.54 | 0.50 | 0.49 | 0.51 | 0.51 |
| **Specificity** | 0.94 | 0.90 | 0.93 | 0.92 | 0.92 | 0.93 |
| **Standardized PPV** | 0.27 | 0.18 | 0.23 | 0.20 | 0.22 | 0.23 |
| **Standardized WDR** | 3.8 | 5.4 | 4.4 | 4.9 | 4.6 | 4.4 |

\*Table adapted from Linnen et al. (1). Since PPV and WDR are highly sensitive to the event rate, we show the standardized event rate at 4% to normalize these values across studies.

##### 3.2.3. Prediction Explanations Increase System Transparency

To explain our classifier’s predictions, we used Shapley Additive explanation (SHAP; version 0.24) [37], [38], and details about this are in **Appendix B.1.3**. SHAP is a tool that ranks features that are most important to a particular prediction. SHAP outputs a Shapley value per feature (feature attribution) for each prediction. Positive values mean the feature is important for predicting the adverse event, and negative values mean the feature is important for predicting no adverse event. The larger the magnitude, the more important the feature was for the class prediction. **Table D.2** shows an example ranked list of explanations for a prediction. **Table 4** shows the most common explanations reported by SHAP for PICTURE predictions within 24 hours of an adverse event. Across all four adverse event types, respiratory rate and shock index were the most common explanations. We also computed the most important features across all predictions to get a global feature importance. This was determined by computing the mean absolute Shapley values across all observations. We found that respiratory rate, time, shock index, blood urea nitrogen, magnesium, minimum oxygen saturation over the last 24 hours, lactate, and heart rate (**Figure D.8**) were globally the most important features. We also considered feature attribution interactions in **Appendix C.2.3**. Therefore, SHAP has the capability of identifying important features for individual predictions, as well as globally across all predictions.

Prediction explanation is essential for system transparency and efficient clinician decision support. Our SHAP prediction explanations have face validity because shock index was a top explanation given for cardiac arrest patients. We are not aware of any other tree-based early warning system that [9], [14] has prediction explanations. Early warning systems have had some challenges for adoption because clinicians have found it difficult to trust ‘black-box’ algorithms[15]. Our utilization of SHAP reduces the ‘black-box’ nature of PICTURE, which should increase the potential for clinicians to trust our system and enhance its value in preventative healthcare.

**Table 4:**
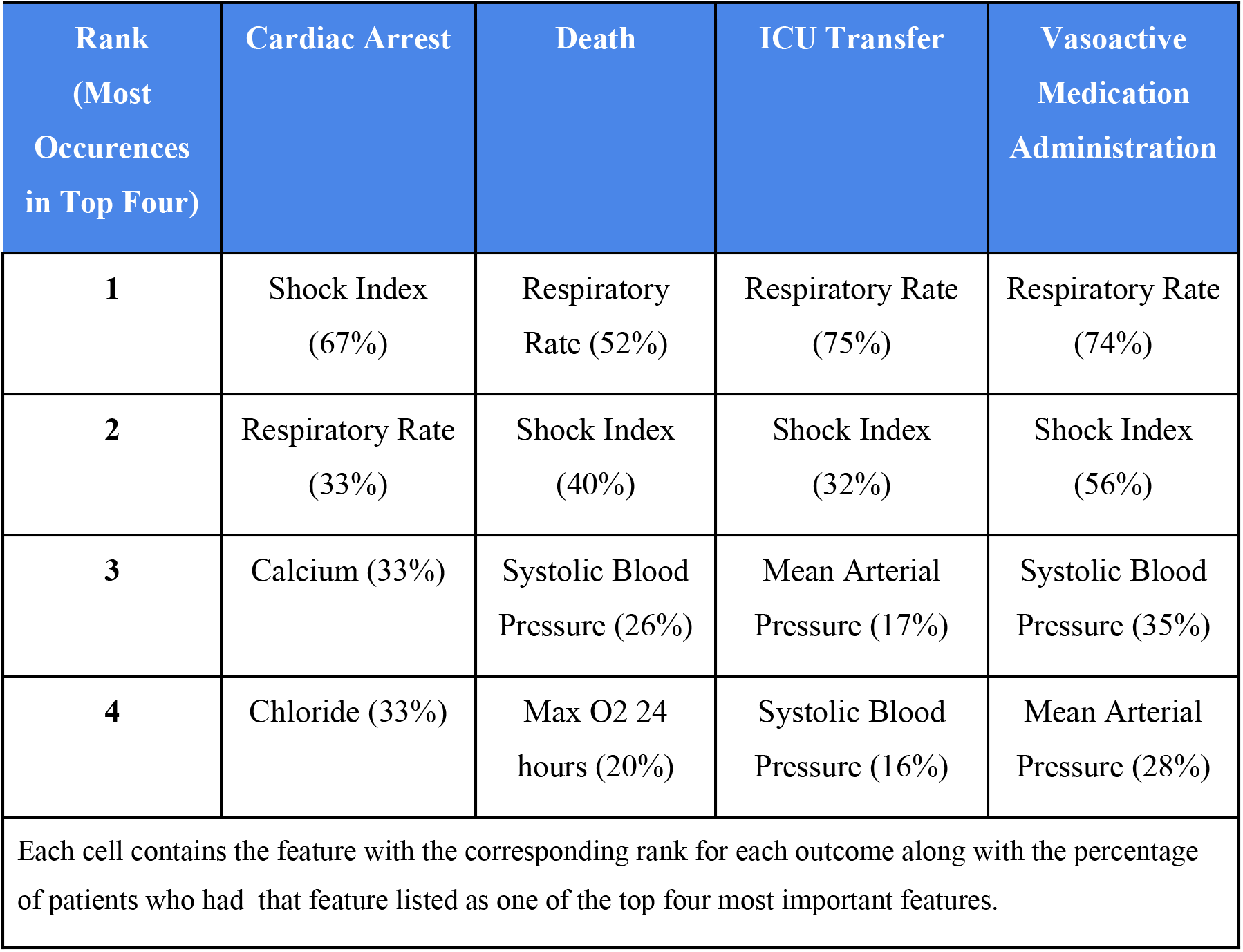
Top four most common features across outcomes as reported by SHAP for correctly predicted events.

| Rank<br>(Most<br>Occurrences<br>in Top Four) | Cardiac Arrest | Death | ICU Transfer | Vasoactive<br>Medication<br>Administration |
| --- | --- | --- | --- | --- |
| 1 | Shock Index<br>(67%) | Respiratory<br>Rate (52%) | Respiratory Rate<br>(75%) | Respiratory Rate<br>(74%) |
| 2 | Respiratory Rate<br>(33%) | Shock Index<br>(40%) | Shock Index<br>(32%) | Shock Index<br>(56%) |
| 3 | Calcium (33%) | Systolic Blood<br>Pressure (26%) | Mean Arterial<br>Pressure (17%) | Systolic Blood<br>Pressure (35%) |
| 4 | Chloride (33%) | Max O2 24<br>hours (20%) | Systolic Blood<br>Pressure (16%) | Mean Arterial<br>Pressure (28%) |
| Each cell contains the feature with the corresponding rank for each outcome along with the percentage of patients who had that feature listed as one of the top four most important features. |  |  |  |  |

#### 3.3. Limitations

The primary limitation of this study is that our cohort was relatively non-diverse and the results are based on a single center. Despite this, PICTURE had consistent performance across demographic subgroups. Also, our simulation results suggest PICTURE will be robust to missingness pattern changes (which may occur when implemented at a different center). In the future, the simulation could be further enhanced by including race and sex; to include these, the loss function would need further enhancements to model categorical, continuous, and binary variables simultaneously. We plan to address this in future work. Additionally, we are actively working toward testing PICTURE at a different hospital.

## 4. CONCLUSION

It is known that early warning systems have challenges generalizing from one hospital to another due to differing population characteristics. However, it is underappreciated that methods used to address missing data during model development also have significant consequences for the generalization of early warning system performance. We demonstrated that tree-based classifiers and logistic regression with indicators can learn missingness patterns in EHR data, and for the first time we have shown that the performance of these models will worsen if the missingness pattern changes. This has significant implications because it helps partially explain why early warning systems developed for preventive healthcare may not be generalizable from hospital to hospital.

To show the existence of the problem and identify its solution, we developed a novel variational autoencoder to generate synthetic EHR data for early warning systems. Our variational autoencoder has a custom loss function specifically designed to operate in high missingness settings characteristic of EHR data. Due to the challenges of comparing machine-learning early warning systems, we suggest our variational autoencoder could be used to facilitate this process. In particular, researchers could generate synthetic data using our autoencoder, which would produce data similar to our hospital’s data, and they could then apply their early warning system to this synthetic data. This could give a common dataset for early warning system comparison. Additionally, if other researchers retrain our variational autoencoder on their hospital’s own data, they could generate synthetic data that could also be released. This could lead to multiple trained variational autoencoders leading to additional comparison datasets. Overall, this synthetic data could be used to improve the training process to enhance all machine-learning early warning system development, which would ultimately improve secondary and tertiary preventive healthcare.

Based on the simulation results, we determined that two imputation methods prevent the missingness pattern from being learned, thus resulting in higher generalization performance: Bayesian regression and randomized imputation approaches. We applied the principles learned from the simulation to develop PICTURE, a tree-based machine learning model intended to aid in preventive healthcare by predicting patient deterioration. PICTURE performed significantly better than common expert-defined aggregate scores (NEWS, SOFA) and logistic regression in predicting general deterioration in this study. PICTURE’s performance is also comparable to state-of-the-art machine learning–based systems. Notably, because we used an appropriate imputation approach based on our simulation results, this prevented PICTURE from learning the missingness patterns, thus increasing its likelihood of providing new information to providers and enhancing its generalizability. Finally, PICTURE is the first tree-based early warning system to add transparency to its predictions using state-of-the-art explanation methods [37], [38] that rank the features most important for each prediction.

## Data Availability

The code for our autoencoder is available here: https://github.com/MCIRCC/picture-simulation

## ACKNOWLEDGEMENTS

The authors acknowledge the University of Michigan Medical School Research Data Warehouse and DataDirect for providing data aggregation, management, and distribution services in support of the research reported in this publication. Kheterpal, S. (2015) Research Data Warehouse/DataDirect: a self-serve tool for data retrieval. Available at the University of Michigan at: https://datadirect.med.umich.edu/. Meagan Ramsey, PhD, provided editing for this manuscript.

## Appendix A.

### Acronyms and Abbreviations

AUROC: Area Under the Receiver Operating Characteristic Curve
AUPR: Area Under The Precision Recall Curve
eCART: Electronic Cardiac Arrest Risk Triage
EHR: Electronic Health Records
ICU: Intensive Care Unit
NEWS: National Early Warning Score
PICTURE: Predicting Intensive Care Transfers and other UnfoReseen Events
PPV: Positive Predictive Value
SOFA: Sequential Organ Failure Assessment Score
SHAP: Shapley Additive explanation
XGBoost: eXtreme Gradient Boosting
WDR: Workup-to-detection ratio

## Appendix B. Supplemental Methods

### B.1. Additional PICTURE Model Development Information

In this section, we discuss methodologic development issues for our PICTURE analytic. Specifically, we discuss how we trained the model using observation encounter data, our imputation approach, and the training algorithm and how we explained our predictions.

#### B.1.1. Windowing to Normalize Training Observations

Patients have different frequencies of laboratory and vital sign orders depending on their level of care and illness severity, so we broke a patient’s time in the hospital into discrete 8-hour intervals (i.e., windows) to train the model (**Figure D.2**). This discrete-time framework [26] normalizes the number of observations for a given amount of time in the hospital. We assigned feature values using the most recent value prior to the start of a window. We allowed a one-hour tolerance for missing features to be observed. Data processing was performed using the Python library *pandas* (version 0.24) [39]. A window was labeled ‘1’ if an adverse event occurred within the window and ‘0’ otherwise. Within a patient encounter, all encounters after the first window labeled ‘1’ were excluded. We are therefore predicting the first adverse event within an encounter.

#### B.1.2. Data Imputation

The EHR has a high percentage of missing data, so we were careful to ensure our tree-based classifier did not learn missing value indicators, which is undesirable (as shown in this paper) because the model may assign more weight to the presence and absence of a variable rather than its value, and performance could degrade if the missingness pattern changed. Based on the results of the simulation (described in **Section 2.7**), we used a Bayesian regression imputation method, which predicts missing values via a regression model using the observed variables. Initially missing variables were imputed using mean imputation to start the iterative process. The imputed value from the previous round was used to predict the missing variables using Bayesian regression. This process was repeated a number of times (20 rounds of iterative refinement).

#### B.1.3. Model Training and Prediction Explanation

We trained the final PICTURE model in Python (version 3.6) using XGBoost [27], an open source library for gradient boosting trees (version 0.90). In brief, gradient boosting tree methods train an ensemble of weak learners in sequence, where the errors of a previous ensemble of trees are iteratively improved until some convergence criteria are met. We optimized hyperparameters (including the max tree depth, the learning rate, and the fraction of data to be sampled with replacement for each tree) using the validation set. We used early stopping to maximize the validation-set AUPR and prevent overfitting. Our training objective function was the binary logistic loss function.

To explain our classifier’s predictions, we used Shapley Additive explanation (SHAP; version 0.24) [37]. SHAP uses Shapley values, a concept from cooperative game theory, to assign feature attribution for particular predictions. Shapley values have consistent and locally accurate feature attribution, unlike other popular prediction explanation methods [37]. SHAP gives explanations consistent with human intuition [38].

SHAP can produce an additive ranked list of features that contribute most to a prediction. We call this additive ranked list for a particular prediction the explanations. SHAP estimates the function *g* that additively assigns feature attributions, *Φ_i_*, particular observation. to each feature in the model for a particular observation.

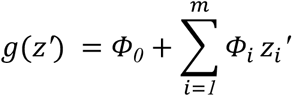

The *Φ_0_* term is the baseline attribution and is defined as the expected prediction score from the model to be explained. The *z_i_* term is 0 if feature *i* is missing and 1 if it is present. For our model, all features are present after imputation. To rank the features most important for a model prediction, we rank the *Φ_i_* terms by their absolute values. Positive *Φ_i_* terms increase the likelihood prediction score, while negative *Φ_i_* terms decrease the prediction score.

SHAP also has the capability to calculate feature interactions, and we explored this aspect as well.

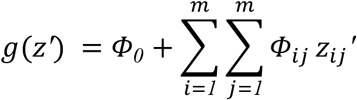

When *i* equals *j*, the *Φ_ii_* terms are defined as the main effects and when *i* is not equal to *j*, the *Φ_ij_* terms are the interaction effects. The additive explanation model is related to the interaction explanation model via 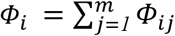.

### B.2. Custom Variational Autoencoder loss function to account for high missing data

We took into consideration two items specific to EHR data in making our model variational autoencoder to simulate realistic data. Specifically, given the high missing rate inherent to the EHR and our desire to simulate both continuous predictors and the binary target variable we had to make some adjustments to standard loss functions for variational autoencoders. First, we needed to specify a loss function to work with continuous and binary variables. Second, due to the high missingness inherent to EHR data, we excluded imputed values in the loss function reconstructions because this would bias the results.

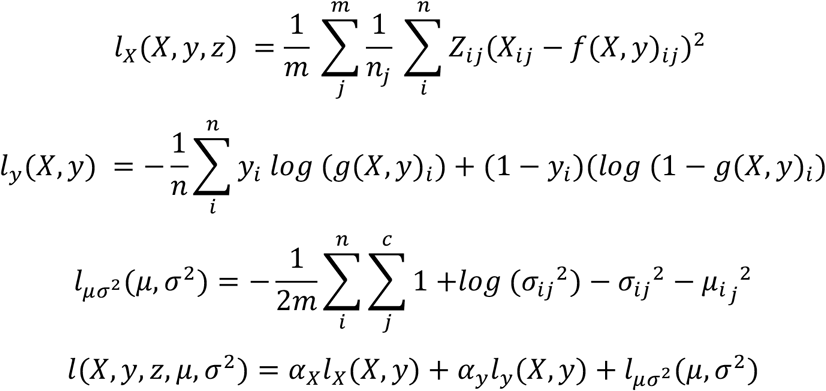

In the above formulation, *X* is the predictor matrix with *n* observations in a batch and *m* is the number of features (predictors) per observation, *Z* is an *n* × *m* indicator matrix whose cells are ‘0’ when the variable was imputed for feature *j* of observation *i* and ‘1’ otherwise, *y* is the target vector with *n* observations, *f*(*X,y*)_*ij*_ is the variational autoencoder output corresponding to the prediction matrix reconstruction for row *i* and column *j* of the predictor matrix, *g*(*X,y*)_*i*_ is the target reconstruction for observation 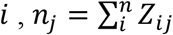 is the number of observed values for feature *j, μ* is an *n* × *c* matrix containing the mean coding vector per observation outputted from the encoder, *σ*^2^ is an *n* × *c* matrix containing the variance coding vector per observation outputted from the encoder, *c* is the coding size, *a_x_* is the hyperparameter weight for *l_x_*(*X,y,z,*), and *a_y_* is the hyperparameter weight for *l_x_*(*X,y*). In words, *l_x_*(*X,y,z,*) is the average of the average reconstruction losses per predictor variable, in terms of mean square error, where for each variable we compute the average reconstruction loss only over observed values. This gives each predictor variable equal weight, preserves the correlation structure, and imputed variables do not contribute to the loss reconstructions. The motivation for this loss function came from comparing the observed correlation matrix to the reconstruction correlation matrix in the training data, and our goal was to make these matrices have a similar structure. The *a_x_* and *a_y_* hyperparameters were selected also by comparing these correlation matrices. For *l_μσ_*^2^(*μ,σ*^2^), we followed the previously published formulations [32].

### B.3. Simulation of learning missingness with a single predictor

The purpose of this second simulation is to show that tree-based models can learn a missingness pattern even when a single predictor is used. This further illuminates how this process occurs so early warning system designers can decide if they should address it.

#### B.3.1. Tree-based classifier imputation

We constructed a single-variable simulation to show that a tree classifier can learn unintended missing data patterns. We compared the performance of three missing data imputation strategies to show that random imputation over the healthy range has superior performance over other tested imputation strategies when the missing pattern changes.

Let Y be a Bernoulli random variable with an event rate of 25%. Y equals to one implies an adverse event happened and Y equals zero implies no adverse event happened. Let X be a random variable whose distribution is conditional on the value of Y. Specifically, let X be distributed normally with a mean equal to zero when Y is zero, and let X be distributed normally with a mean equal to one when Y is one. The standard deviation of X was fixed at one. We then fixed the missing rate of X to be 10% when Y is one. We varied the missing rate of X from 10% to 90% when Y is equal to zero. When the difference of missing rates of X for each class (Y equals to 0 and Y equals to one) is large, this models a situation similar to the one we observe in our data for adverse events (Y) and lactate (X). A lactate test may be ordered when a physician suspects an adverse event will occur. This results in a lower missing rate for patients with an adverse event.

We constructed four tree classifiers to demonstrate the consequences of different imputation strategies when the missingness pattern changes. In particular, we investigated how the classifiers performed at test time when the data became completely observed. In other words, we trained these models in situations with missing data as described above and tested in an environment with no missing data. We tested a decision tree using fully observed data, a decision tree with mean imputation of healthy values, a decision tree with extreme value imputation, and a decision tree with random healthy value imputation. We used 40,000 samples for training and 10,000 samples for testing the models.

#### B.3.2. Rationale for a single predictor imputation strategy

We have designed PICTURE to make predictions based on the values for each test, not factoring in the significance of whether or not a variable was observed. Although missing labs do correlate with the patient’s condition, exactly which labs are missing is susceptible to change for a number of reasons. For example, changes in hospital policy or protocols can affect which labs are routinely taken. Attitudes of clinicians toward testing, level of clinical training and experience, or personal preference of individual practitioners can also affect which labs are ordered, regardless of the patient’s condition. If we had allowed PICTURE to factor in which labs were taken, this would create issues because if the missing pattern of labs changed for any reason, the efficacy of PICTURE’s predictions would decline because it learned to make its prediction based on the presence or absence of a lab and not its value. Our imputation strategy solves this problem.

To further illustrate this, consider that certain labs (like lactate) are commonly only taken when a patient has a severe condition, thus only the sicker patients undergo such lab testing. If missing value indicators are included in predictive power, ordering those labs would alarm that the patient’s condition is worsening, regardless of the lab results. The outcome of this would be that a patient getting an uncommon lab would appear sick even if the lab value was normal. Similarly, there would be little change in the deterioration score of a patient who had an abnormal value for an infrequent test that later returned to normal. This is because the presence of the labs being taken are correlated with sicker patients. Furthermore, this would be confusing to clinicians who would see the infrequently ordered labs as the reason for an increased risk score, even if those values returned to normal. For example, sicker patients tend to have lactate tested while healthier patients do not. This presents itself as a problem at runtime. The model realizes that patients tested for lactate are in a more serious condition. The clinicians would only see that lactate seems to be a useful test value. If they start ordering it because of its importance to the model, PICTURE would start to flag more patients as deteriorating. The lactate lab in turn could raise false alarms on patients even if their labs came back normal.

## Appendix C. Supplemental Results and Discussion

### C.1. Tree-Based Models can Learn Missingness Patterns Even with a Single Predictor

We also investigated if this learning of the missingness pattern could be observed using a single variable. The purpose is to understand the ease with which this problem could occur and further show its effects on generalizability performance. To assess this, we implemented a separate small simulation analysis to illustrate that tree-based methods can learn missingness patterns even with a single variable (**Table D.3**). In the most extreme case, when the difference in missing rate was 0.8, the “Random Healthy” imputation prevented the missing indicator pattern from being learned and experienced a drop in PPV of 0.01, the “Mean” imputation had a drop of 0.47, and the “Extreme Value” imputation approach had a drop in PPV of 0.50. **Figures D.9 and D.10** show the results of a sensitivity analysis for how the performance drops as the missing rate between classes (Y = 0 [those with an adverse event] and Y = 1 [those without an adverse event]) changes. The primary conclusion is: as the difference in missing rate between classes increases, performance worsens more for “Mean” and “Extreme Value” imputation approaches.

These results underscore the importance of carefully imputing data for early warning systems using tree-based models. They also highlight that this problem occurs when a variable has different missing rates in patients who do have events versus those who do not have events. This could be due to clinicians ordering more labs (e.g., lactate) when a patient has a more severe illness, which underscores that predictions should be based on the test values themselves, not the presence or absence of lab values. By addressing this, we are theoretically showing that PICTURE is robust to missingness pattern changes. Thus, we expect PICTURE will continue to make accurate predictions if hospital policy or physician behavior changes which tests are routinely ordered.

### C.2. Additional PICTURE Analyses

In these sections, we analyze how early PICTURE makes a prediction in the lead time analysis, we assess how well PICTURE performs in subgroups within our cohort, and we analyze feature attribution interactions for prediction explanations. Both of these are necessary to understand the clinical impact of our tool.

#### C.2.1. Lead Time Analysis: PICTURE Predicts Early Enough for Possible Intervention

We performed a lead time analysis, selecting PICTURE’s threshold such that its encounter-level sensitivity was 51%. To compute the lead time at this threshold, we selected the patients who would have alarmed at this threshold and selected the earliest time point at which the alarm would fire. At this threshold, PICTURE had a PPV of 0.26 (3.4% event rate), a WDR of 4, and a median lead time of alarming 23 hours before the adverse event (the 25-percentile lead time was 3 hours, and the 75-percentile lead time was 85 hours). Rapid response teams calls may precede an adverse event. If PICTURE could predict these rapid response team calls before they occurred, then this result may suggest that PICTURE is providing new information to clinicians when it sends an alert. We found that PICTURE predicted rapid response team events preceding adverse events with a median lead time of 24 hours. These results suggest that PICTURE could have alerted the rapid response team before they were called to respond to potential patient deterioration events.

#### C.2.2. PICTURE Performed Similarly Across Subgroups

When predicting deterioration events, the PICTURE AUROC was consistent across different patient strata, including sex, race, and other outcomes (**Table D.4**). PICTURE predicted death with a much higher AUROC than other events. Since cardiac arrest had such a low event rate, the raw AUPR was low. Part of the reason for lower performance when predicting cardiac arrest could be due to how events were labeled in our cohort. Encounters were grouped into event groups based on the earliest event per encounter. Additionally, all observations after the first event were removed from further analysis because we are predicting the first deterioration event. For example, if a patient received vasoactive medications prior to the cardiac arrest, the patient would be classified under the vasoactive subgroup.

#### C.2.3. Feature Attribution Interactions Are Less Substantial Than Main Feature Attributions

We also considered SHAP’s interaction feature attributions for further enhancing prediction explanations. However, for our model, we found the interaction feature attributions to be much smaller than the main feature attributions. Specifically, when we looked at the maximum interaction feature attributions across the 1% highest model prediction scores, we found that the feature attribution score at 90%-tile for interaction feature attributions was 0.097, while 15%-tile for main feature attributions was 0.11. This indicates the main feature attributions are much larger in magnitude than the interaction feature attributions and therefore explain the predictions more than the interaction features.

## Appendix D. Supplemental Tables and Figures

Please see attached supplemental file.

## REFERENCES

[1] D. Linnen, G. J. Escobar, X. Hu, E. Scruth, V. Liu, and C. Stephens, “Statistical Modeling and Aggregate-Weighted Scoring Systems in Prediction of Mortality and ICU Transfer: A Systematic Review,” J. Hosp. Med., vol. 14, no. 3, pp. 161–161, Mar. 2019, doi: 10.12788/jhm.3151.

[2] S. R. Bapoje, J. L. Gaudiani, V. Narayanan, and R. K. Albert, “Unplanned transfers to a medical intensive care unit: causes and relationship to preventable errors in care,” J. Hosp. Med., vol. 6, no. 2, pp. 68–72, Feb. 2011, doi: 10.1002/jhm.812.

[3] D. R. Levinson, “ADVERSE EVENTS IN HOSPITALS: NATIONAL INCIDENCE AMONG MEDICARE BENEFICIARIES,” 2010.

[4] M. D. Le Lagadec and T. Dwyer, “Scoping review: The use of early warning systems for the identification of in-hospital patients at risk of deterioration,” Aust. Crit. Care Off. J. Confed. Aust. Crit. Care Nurses, vol. 30, no. 4, pp. 211–218, Jul. 2017, doi: 10.1016/j.aucc.2016.10.003.

[5] J. McGaughey, F. Alderdice, R. Fowler, A. Kapila, A. Mayhew, and M. Moutray, “Outreach and Early Warning Systems (EWS) for the prevention of intensive care admission and death of critically ill adult patients on general hospital wards,” Cochrane Database Syst. Rev., no. 3, p. CD005529, Jul. 2007, doi: 10.1002/14651858.CD005529.pub2.

[6] J. A. Kempker, H. E. Wang, and G. S. Martin, “Sepsis is a preventable public health problem,” Crit. Care, vol. 22, no. 1, p. 116, May 2018, doi: 10.1186/s13054-018-2048-3.

[7] M. P. Young, V. J. Gooder, K. McBride, B. James, and E. S. Fisher, “Inpatient Transfers to the Intensive Care Unit,” J. Gen. Intern. Med., vol. 18, no. 2, pp. 77–83, Feb. 2003, doi: 10.1046/j.1525-1497.2003.20441.x.

[8] M. J. Rothman, “The Emperor Has No Clothes,” Crit. Care Med., vol. 47, no. 1, pp. 129–130, Jan. 2019, doi: 10.1097/CCM.0000000000003505.

[9] M. M. Churpek, T. C. Yuen, C. Winslow, D. O. Meltzer, M. W. Kattan, and D. P. Edelson, “Multicenter Comparison of Machine Learning Methods and Conventional Regression for Predicting Clinical Deterioration on the Wards,” Crit. Care Med., vol. 44, no. 2, pp. 368–374, Feb. 2016, doi: 10.1097/CCM.0000000000001571.

[10] P. Kipnis et al., “Development and validation of an electronic medical record-based alert score for detection of inpatient deterioration outside the ICU,” J. Biomed. Inform., vol. 64, pp. 10–19, Dec. 2016, doi: 10.1016/j.jbi.2016.09.013.

[11] T. Desautels et al., “Using Transfer Learning for Improved Mortality Prediction in a Data-Scarce Hospital Setting,” Biomed. Inform. Insights, vol. 9, pp. 117822261771299–117822261771299, Jan. 2017, doi: 10.1177/1178222617712994.

[12] T. Desautels et al., “Prediction of early unplanned intensive care unit readmission in a UK tertiary care hospital: a cross-sectional machine learning approach.,” BMJ Open, vol. 7, no. 9, pp. e017199–e017199, Sep. 2017, doi: 10.1136/bmjopen-2017-017199.

[13] C. A. Alvarez et al., “Predicting out of intensive care unit cardiopulmonary arrest or death using electronic medical record data,” BMC Med. Inform. Decis. Mak., vol. 13, p. 28, Feb. 2013, doi: 10.1186/1472-6947-13-28.

[14] M. Green, H. Lander, A. Snyder, P. Hudson, M. Churpek, and D. Edelson, “Comparison of the Between the Flags calling criteria to the MEWS, NEWS and the electronic Cardiac Arrest Risk Triage (eCART) score for the identification of deteriorating ward patients,” Resuscitation, vol. 123, pp. 86–91, Feb. 2018, doi: 10.1016/j.resuscitation.2017.10.028.

[15] H. M. Giannini et al., “A Machine Learning Algorithm to Predict Severe Sepsis and Septic Shock: Development, Implementation, and Impact on Clinical Practice*,” Crit. Care Med., vol. 47, no. 11, p. 1485, Nov. 2019, doi: 10.1097/CCM.0000000000003891.

[16] L.-H. Fu et al., “Development and validation of early warning score system: A systematic literature review,” J. Biomed. Inform., vol. 105, p. 103410, May 2020, doi: 10.1016/j.jbi.2020.103410.

[17] A. Sharafoddini, J. A. Dubin, D. M. Maslove, and J. Lee, “A New Insight Into Missing Data in Intensive Care Unit Patient Profiles: Observational Study,” JMIR Med. Inform., vol. 7, no. 1, p. e11605, Jan. 2019, doi: 10.2196/11605.

[18] E. Ford, P. Rooney, P. Hurley, S. Oliver, S. Bremner, and J. Cassell, “Can the Use of Bayesian Analysis Methods Correct for Incompleteness in Electronic Health Records Diagnosis Data? Development of a Novel Method Using Simulated and Real-Life Clinical Data,” Front. Public Health, vol. 8, p. 54, 2020, doi: 10.3389/fpubh.2020.00054.

[19] J. Yoon, L. N. Drumright, and M. Van Der Schaar, “Anonymization through Data Synthesis using Generative Adversarial Networks (ADS-GAN),” IEEE J. Biomed. Health Inform., Mar. 2020, doi: 10.1109/JBHI.2020.2980262.

[20] A. Reiner Benaim et al., “Analyzing Medical Research Results Based on Synthetic Data and Their Relation to Real Data Results: Systematic Comparison From Five Observational Studies,” JMIR Med. Inform., vol. 8, no. 2, p. e16492, Feb. 2020, doi: 10.2196/16492.

[21] A. Tuladhar, S. Gill, Z. Ismail, and N. D. Forkert, “Building machine learning models without sharing patient data: A simulation-based analysis of distributed learning by ensembling,” J. Biomed. Inform., vol. 106, p. 103424, Jun. 2020, doi: 10.1016/j.jbi.2020.103424.

[22] M. K. Baowaly, C. C. Lin, C. L. Liu, and K. T. Chen, “Synthesizing electronic health records using improved generative adversarial networks,” J. Am. Med. Inform. Assoc. JAMIA, vol. 26, no. 3, pp. 228–241, 01 2019, doi: 10.1093/jamia/ocy142.

[23] D. P. Kingma and M. Welling, “Auto-Encoding Variational Bayes,” ArXiv13126114 Cs Stat, Dec. 2013, Accessed: Oct. 07, 2019. [Online]. Available: http://arxiv.org/abs/1312.6114.

[24] B. D. Ripley, Pattern Recognition and Neural Networks. Cambridge: Cambridge University Press, 1996.

[25] P. Rajpurkar et al., “Deep learning for chest radiograph diagnosis: A retrospective comparison of the CheXNeXt algorithm to practicing radiologists,” PLOS Med., vol. 15, no. 11, p. e1002686, Nov. 2018, doi: 10.1371/journal.pmed.1002686.

[26] M. M. Churpek et al., “Multicenter Development and Validation of a Risk Stratification Tool for Ward Patients,” Am. J. Respir. Crit. Care Med., vol. 190, no. 6, pp. 649–655, Sep. 2014, doi: 10.1164/rccm.201406-1022OC.

[27] T. Chen and C. Guestrin, “XGBoost: A Scalable Tree Boosting System,” in Proceedings of the 22Nd ACM SIGKDD International Conference on Knowledge Discovery and Data Mining, New York, NY, USA, 2016, pp. 785–794, doi: 10.1145/2939672.2939785.

[28] G. B. Smith, D. R. Prytherch, P. Meredith, P. E. Schmidt, and P. I. Featherstone, “The ability of the National Early Warning Score (NEWS) to discriminate patients at risk of early cardiac arrest, unanticipated intensive care unit admission, and death,” Resuscitation, vol. 84, no. 4, pp. 465–470, Apr. 2013, doi: 10.1016/j.resuscitation.2012.12.016.

[29] Royal College of Physicians, “National Early Warning Score (NEWS): Standardising the assessment of acute illness severity in the NHS.” Royal College of Physicians, London, 2012.

[30] J. L. Vincent et al., “The SOFA (Sepsis-related Organ Failure Assessment) score to describe organ dysfunction/failure. On behalf of the Working Group on Sepsis-Related Problems of the European Society of Intensive Care Medicine,” Intensive Care Med., vol. 22, no. 7, pp. 707–710, Jul. 1996.

[31] S. Yu et al., “Comparison of risk prediction scoring systems for ward patients: a retrospective nested case-control study,” Crit. Care, vol. 18, no. 3, p. R132, Jun. 2014, doi: 10.1186/cc13947.

[32] Géron, Aurélien, Hands-On Machine Learning with Scikit-Learn, Keras, and TensorFlow, 2nd ed. O’Reilly Media, Inc., 2019.

[33] Klambauer, Günter, Unterthiner, Thomas, Mayr, Andreas, and Hochreiter, Sepp, “Self-Normalizing Neural Networks,” in Proceedings of the 31st International Conference on Neural Information Processing Systems, 2017, pp. 972–981.

[34] J. Ma, Q. Zhang, J. Lou, J. C. Ho, L. Xiong, and X. Jiang, “Privacy-Preserving Tensor Factorization for Collaborative Health Data Analysis,” Proc. ACM Int. Conf. Inf. Knowl. Manag. ACM Int. Conf. Inf. Knowl. Manag., vol. 2019, pp. 1291–1300, Nov. 2019, doi: 10.1145/3357384.3357878.

[35] R. Barth, J. IJsselmuiden, J. Hemming, and E. J. Van Henten, “Synthetic bootstrapping of convolutional neural networks for semantic plant part segmentation,” Comput. Electron. Agric., vol. 161, pp. 291–304, Jun. 2019, doi: 10.1016/j.compag.2017.11.040.

[36] B. Moiseev, A. Konev, A. Chigorin, and A. Konushin, “Evaluation of Traffic Sign Recognition Methods Trained on Synthetically Generated Data,” in Advanced Concepts for Intelligent Vision Systems, Cham, 2013, pp. 576–583, doi: 10.1007/978-3-319-02895-8_52.

[37] S. M. Lundberg, G. G. Erion, and S. I. Lee, “Consistent Individualized Feature Attribution for Tree Ensembles,” ArXiv180203888 Cs Stat, Feb. 2018, Accessed: Mar. 27, 2019. [Online]. Available: http://arxiv.org/abs/1802.03888.

[38] S. M. Lundberg and S. I. Lee, “A Unified Approach to Interpreting Model Predictions,” in Advances in Neural Information Processing Systems 30, I. Guyon, U. V. Luxburg, S. Bengio, H. Wallach, R. Fergus, S. Vishwanathan, and R. Garnett, Eds. Curran Associates, Inc., 2017, pp. 4765–4774.

[39] W. McKinney, “Data Structures for Statistical Computing in Python,” presented at the Proceedings of the 9th Python in Science Conference, 2010, pp. 51–56, Accessed: Jul. 12, 2019. [Online]. Available: http://conference.scipy.org/proceedings/scipy2010/mckinney.html.

